# Point-of-Care Solid-Phase PCR in Vertical Microfluidic Chip Integrated with All-Dielectric Nanostructured Metasurface for Highly -Sensitive, Multiplexed Pathogen Detection

**DOI:** 10.1101/2025.05.24.25328289

**Authors:** Seder Fayyad, Leonid Yu. Beliaev, Rodrigo Coronel Téllez, Christian Anthon, Dhouha Grissa, Tao Zheng, Jan Gorodkin, Sanshui Xiao, Yi Sun

**Affiliations:** Department of Health Technology, Technical University of Denmark, Ørsteds Plads, DK-2800 Kgs. Lyngby, Denmark; Department of Electrical and Photonics Engineering, Technical University of Denmark, Ørsteds Plads, DK-2800 Kgs. Lyngby, Denmark; Center for non-coding RNA in Technology and Health, Department of Veterinary and Animal Sciences, Faculty of Health and Medical Sciences, University of Copenhagen, Frederiksberg, Denmark; NanoPhoton—Center for Nanophotonics, Technical University of Denmark, Building 345A, DK-2800 Kongens Lyngby, Denmark

**Keywords:** Multiplex detection, dielectric nanostructure, vertical microfluidic

## Abstract

Multiplexed solid-phase polymerase chain reaction (SP-PCR) has emerged as an indispensable modality for concurrent amplification of multiple genetic loci within a singular reaction vessel, facilitating efficient molecular diagnostics. Nevertheless, SP-PCR has seldom been integrated into point-of-care diagnostic devices due to several technical challenges, such as bubble formation during PCR, long reaction time and low fluorescence signals generated from the PCR products on a solid surface. To circumvent these constraints, we engineered a microfluidic chip comprising SP-PCR and nanophotonic enhancement to enable highly sensitive, high-throughput, and cost-efficient molecular diagnostics. The chip’s vertical orientation integrates preloaded reagent chambers for sequential lysis, washing, elution, and amplification, driven by a synchronized stepper motor and air vacuum, achieving robust nucleic acid purification and reverse transcription-PCR and enables bubble-free, gravity-assisted fluid dynamics during the PCR thermocycling. Thermal cycling is expedited through a dual-heater configuration alternating at sub-second intervals, obviating active cooling and shortening the reaction time. All-dielectric nanostructured metasurface was incorporated beneath the PCR chamber, allowing for the facile immobilization of DNA arrays to conduct SP-PCR. Taking advantage of guided-mode resonance supported by the metasurface and the SP-PCR approaches permits multiplexed detection and achieves a detection limit of 10 copies/reaction for SARS-CoV-2, highlighting the platform’s potential for point-of-care diagnostics, personalized medicine, and high-throughput pathogen surveillance. Facile fabrication and automation emphasize scalability for mass production and deployment and collectively represent advancement in point-of-care diagnostics.

## 1. Introduction

Multiplexed solid-phase polymerase chain reaction (SP-PCR) is a powerful molecular biology technique that allows the simultaneous amplification of multiple target DNA sequences within a single reaction and a single-wavelength fluorophore.^1–4^ SP-PCR relies on a DNA array immobilized on a solid substrate comprising of multiple primers, each specific to a different target, enabling the efficient detection and differentiation of various genetic markers or pathogens in a single test. The ability to amplify several targets in one reaction significantly enhances the efficiency of molecular diagnostics, reduces reagent consumption, and minimizes the time required for analysis.^5^ In events like epidemics and infectious disease outbreaks, SP-PCR can play an extensive role in providing the correct assessment and distinction between the seasonal infectious disease and new mutant virus, hence preventing disease widespread.^6^ With the feature of SP-PCR of spatially localized DNA synthesis on functionalized surfaces within a confined reaction microenvironment, minimizing reagent consumption and cross-contamination, SP-PCR appears to synergize seamlessly with microfluidic system.

However, SP-PCR has seldom been integrated into point-of-care diagnostic devices due to several technical challenges of special requirements for liquid assay behavior and low-sensitivity assay, hindering a wide adaptation for such a molecular approach.^7^ For liquid assay behavior, bubble formation within microfluidic PCR chamber disrupts thermal and physical contact of PCR reagent, inheriting a low reaction efficiency that lengthens assay times and yields weak fluorescence signals and it provides unreliable and unrepeated diagnostic results.^8,9^ For example, studies have reported that even microliter-scale gas pockets in PCR chips can expand during thermal cycling, causing partial chamber drying, inconsistent amplification, and spurious signal dropout.^10^ To address such a bubble issue, multi-layer PDMS/glass stacks with on-chip valves,^11^ oil layer^12^ and pressure-control plumbing^13^ merely to purge or isolate bubbles, adding fabrication complexity and cost. On the other hand, SP-PCR often exhibits lower efficiency than conventional liquid PCR because the reaction relies solely on diffusion to bring free primers in the solution into contact with immobilized probes on a solid surface. Extending the annealing and extension steps can partially offset the low efficiency limitation.^14^ However, it also prolongs the overall assay time, exacerbates reagent evaporation and, importantly, reduces the specificity due to an increase in mis-priming and/or it amplifies low-abundance, off-target amplicons.

Enhancing spatially localized electromagnetic fields has been widely explored, primarily through plasmonic nanostructures and photonic microcavities, which focus light at the nanoscale to boost fluorescence detection sensitivity, particularly for low analyte concentrations.^15,16^ However, this method is effective only when the analyte is in close proximity to the region of the strongly confined electromagnetic field. All-dielectric metasurfaces have recently emerged as an attractive alternative for fluorescence enhancement.^17,18^ These metasurface platforms can support guided-mode resonances, achieving comparable field enhancement and large overlap with the analyte, while offering superior spectral control, scalable fabrication, and compatibility with the standard nanofabrication technologies.^17,18^ The combination of field strength and spatial reach makes such guided-mode resonances especially well-suited for biosensing applications. Dielectric metasurfaces based on materials such as silicon,^19^ titanium dioxide,^20^ and silicon nitride^21^ have demonstrated fluorescence-enhancement factors suitable for single-molecule sensitivity in biosensing applications. To date, only one report has integrated an all-dielectric metasurface to read the fluorescence signal of amplified PCR products.^22^ The setup detects an impressive level of sensitivity as low as 1 copy per test. However, the test required a separate bench-top amplification prior to loading the sample above the nanostructure for reading; meanwhile, an expensive electron-beam lithography was used to fabricate such a structure, factors that impede integration into a true lab-on-a-chip and large-scale manufacture.

In this study, we address these gaps by developing an all-in monolithic, wafer-scale fabricated dielectric metasurface device integrated on bubble-free microfluidic chip for multiplexed SP-PCR. The propped chip consists of a vertical microfluidic architecture, wherein the PCR mixture naturally resides at the bottom of PCR chamber, preventing bubble formation and liquid displacement from the solid surface. At the bottom of PCR chamber, we integrated a customized array of metasurface nanostructures, wherein probes for several pathogen targets are immobilized, and later DNA is amplified. To achieve a sample-to-answer system, we also integrate a pre-application step (Nucleic acid purification) with a simplified reagent manipulation technique to handle the fluid addition and trapping via an automated valveless rotating disc (Figure 1, Figure S1 and Video S1A). The vertically oriented PCR chamber geometry ensures that the reaction mixture remains in continuous contact with the functionalized surface, eliminating bubble formation without external valves or pressure control, while the embodied metasurface–SP-PCR module provides a signal-to-background ratio enhancement of ~6 times compared to the flat surface. Such enhancement is essential for patient samples with low-copy targets (<1000 copies). As an automated reagent-handling, self-contained, sample-to-answer and coupled with a metasurface–SP-PCR system, we demonstrate multiplexed detection of viral targets down to 10 copies per reaction (100 % specificity) under a single excitation–emission wavelength, validating the platform’s potential for rapid, reliable, and scalable point-of-care diagnostics.

**Figure 1.**
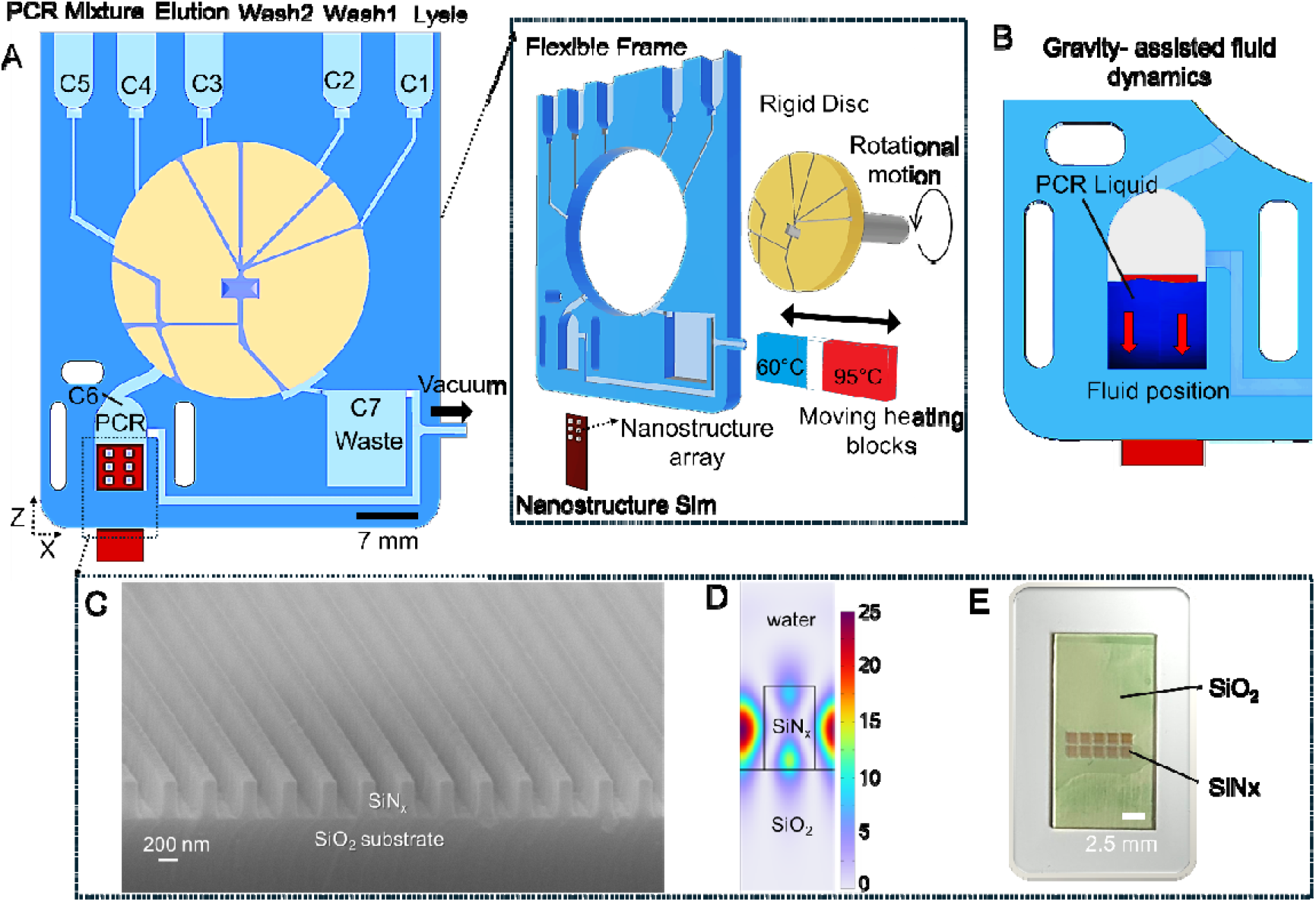
Multiplexed detection system on a vertical chip. (A) Side view of the Assembled chip including an outer frame, an inner disc, and a nanostructure sim for solid phase PCR. The inset shows the disassembled parts of the system prior to the assembly, including two heaters with different temperatures that move alternately behind the PCR chamber. (B) Position of the PCR reagent. Naturally, the reagent remains at the bottom of the chamber, hence a continuous submersion of the solid array during the PCR process. (C) Scanning electron microscope (SEM) cross-sectional images of the fabricated silicon nitride (SiNx) gratings on top of silicon dioxide (SiO2) substrate. The nanostructure is inserted under the PCR chamber with different viral probes to enhance the fluorescence reading. (D) Normalized electric field distributions at the guide-mode resonance of 639 nm. (E). Real image of the SiN_x_ nanostructure on SiO_2_ substrate.

## 2. Methods

### 2.1 Chip fabrication

The disposable chip consists of two parts: 3D-printed inner rigid disc (Ø = 28.35 mm) (Form 3, Biomid resin, Formlabs. Inc) and 3D-printed flexible outer frame part (Form 3 printer, A50 flexible biomed resin, Formlabs. Inc.). The Autodesk software was used to design the structure prior to the 3D-printing. After printing, the 3D-parts soaked in IPA for 20 min then cured in UV light machine for 2 hours and finally washed by DI water 3 times prior to use (Fabrication details in Figure S2). Both inner and outer parts were 3D-printed in as a single-layer print, hence there was no need to close the microchannels. The detailed dimensions of the chip and the microchannel are listed in Figure S3A. The inner rigid disc then inserted manually into the outer flexible frame without the need for special equipment or bonding (Video S1B). The outer frame has 7 chambers for the assay buffer including: sample / lysis chamber 1 (C1), wash 1 (C2), wash buffer 2 (C3), elution buffer (C4), RT-RPA mixture (C5), PCR amplification (C6) and waste chamber (C7). The capacity of the chambers from 1 to 5 are 100μL, although the user can fill them as the assay requires. The silicon nitride sim (10 × 20 mm^2^), which contains nanostructure features, is inserted at the bottom of C6, as a sim, and glued from sidewalls of C6. To rotate the inner disc, a cross-shaped hole is designed in the middle of the inner disc allowing the external motor arm with a cross-shaped shape to engage with the chip (Figure S3B and Video S1C). A hole of 6×4 mm^2^ silica matrix (Y-SM-BC-1, Biocomma) was embedded within the inner rotating disc and then closed with 1 mm 3D-printed layer. The matrix serves as the solid-phase extraction medium for RNA capture (Figure S3C).

### 2.2 Numerical simulation for Nanostructure

Simulations based on the finite element method (FEM) were performed using COMSOL Multiphysics 6.2 (COMSOL AB, Sweden). A periodic nanostructure typically supports multiple guided-mode resonances. To enhance the fluorescence signal, we need to select a resonant mode that strongly overlaps with the region where the fluorophore is located. We chose the guided-mode resonance by solving eigenmodes of the SiNx subwavelength grating and evaluating the mode profiles with a large ratio of energy overlapping with the fluorophore.

The nanostructure geometry was further optimized to align the guided-mode resonance with the Cy5 emission spectrum. We used a refractive index of SiO2 (n = 1.45) and SiNx (n = 2.15) as a result of ellipsometry measurements.^23^ Based on our analysis and optimization, the grating height, period, and bar width were chosen as 359, 410, and 205 nm, respectively.

### 2.3 Nanofabrication of Nanostructure

The fabrication process of the NS was conducted in a class 10–100 cleanroom facility. Initially, 500 μm-thick fused silica wafers underwent standard RCA cleaning. Next, a Si-rich silicon nitride (SiNx) layer, was deposited using low-pressure chemical vapor deposition (LPCVD) (furnace from Tempress). This material was chosen because it has minimal absorption at the wavelengths of interest and a sufficiently high refractive index, allowing for high contrast with the substrate, which is necessary for better field localization (Table S1). The subwavelength grating structures, with a pitch of 410 nm and bar width of 205 nm, were then patterned via large-area deep ultraviolet (DUV) lithography (Canon FPA-3000 EX4, Canon), followed by dry etching (DRIE Pegasus, SPTS Technologies Ltd.). More details can be found in our previous work,^23^ where a similar structure was fabricated. The resulting structures were analyzed using scanning electron microscopy (SEM Zeiss Supra 40VP, Zeiss), as illustrated in Figure 2A.

**Figure 2.**
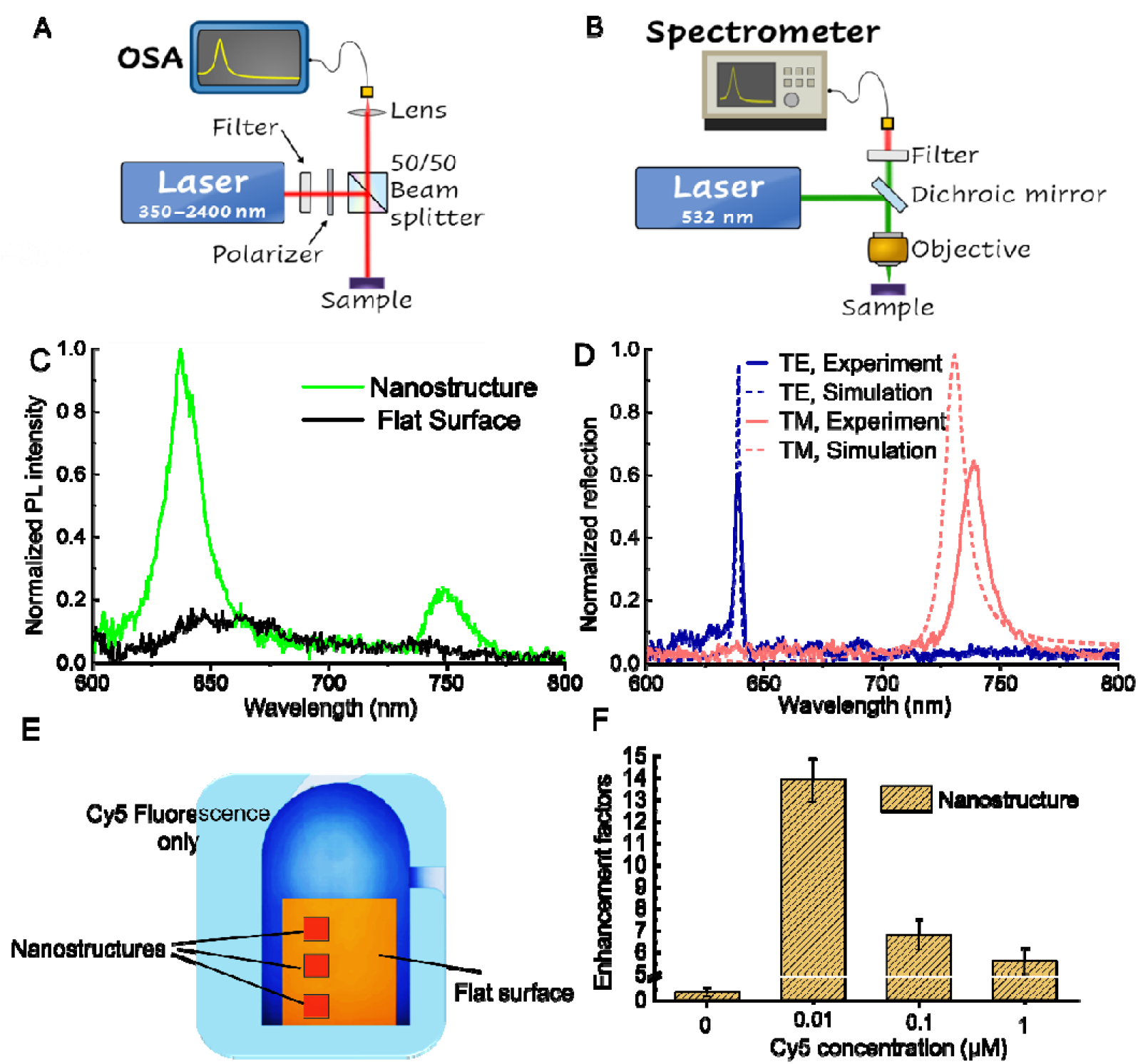
Nanostructure characterization. (A) The optical setup for reflectance measurements. (B) The optical setup for PL measurements. (C) Measured PL spectra of the patterned nanostructures and unpatterned/flat area on the same chip. (D) Reflection spectra of the structure for TE- (blue curve) and TM- (red curve) polarizations in water solution. The presented experimental data (solid lines) and those obtained by numerical simulations (dashed lines). (E-F) The optical performance of the flat surface compared to the nanostructure (NS) surface under Cy5 fluorophore. Background-corrected intensities were obtained by subtracting the autofluorescence of each substrate, and the enhancement factor was determined as the ratio of the net fluorescence intensity from the patterned surface to that from the flat surface.

### 2.4 Fluid control and heating system

The system has three stepper motors (SM) to operate a 3 ml syringe to create vacuum in the chip (SM1), to rotate the inner disc (SM2) and alternate the position of two heaters or creating a shaking motion for the fluid in the amplification chamber (SM3) (Figure S1). The motion of the three motors is preprogrammed and controlled by an Arduino controller circuit (Figure S4). The linear motion of SM2 is changed according to the required flowrate for each buffer addition. On top of SM3, two customized heaters were placed beside each other with a 20 mm distance to provide two different temperatures at the same time (details of the heater dimension in Figure S1). The temperature was controlled using an Arduino board (Arduino Uno Rev3, Arduino). Programming the forward and backward motion of SM3 at high speed (200 mm/s) for short distance (0.5 mm) creates an ideal shaking device for mixing the reagent (Video S1D and E).

### 2.5. One-step RT-RPA reagents and Primer and probe design

Twist Bioscience supplied synthetic RNA standards for SARS⍰CoV⍰2 (MN908947.3), Influenza A H1N1 (NC_026433) and Influenza B (NC_002209). The RNA purification kit was purchased from (QIAamp Viral TNA Min Kit, QIAGEN). For a 40 µL one⍰step RT⍰PCR assay, we used the SensiFAST™ Probe No⍰ROX One⍰Step kit (Meridian Bioscience). Reactions contained 32 µL of master mix, including 12 µL of purified RNA, and 8 µL of a primer mix with all ten forward and reverse primers at 0.8 µM each (Table S2). First, Reverse transcription was performed at 45 °C in C6, followed by a 1-minute polymerase-activation step at 95 °C. Each PCR cycle consisted of an 8 s denaturation at 95 °C and a 22 s for annealing/extension at 60 °C. Forward primers carried a 5′ Cy⍰5 fluorophore for fluorescence detection, while probes featured a 5′ C6⍰amino linker to enable covalent attachment to silicon nitride surfaces. All oligonucleotides were ordered from TAG Copenhagen A/S (Denmark); probes were sourced from Integrated DNA Technologies (Denmark). The primer and probe designs used in this study appear in Table S3.

### 2.6 Optical Characterization

For photoluminescence (PL) measurements, a home-built setup was used (Figure 2B). A 532-nm laser (LLS-0532, Laserglow Technologies) serves as the excitation light source. The incident power was adjusted using a gradient filter (NDC-50C-4M-A, Thorlabs) to 1 mW. The objective lens was Olympus MPLFN50X (50x, NA=0.5). A 600 nm long-pass filter (FELH0600, Thorlabs) is used to filter the pump laser. The PL signals were detected by the spectrometer (ANDOR, SR-303I-A). For the liquid phase PCR with SYBR Green fluorescence, 490 nm and 515 nm excitation and emission filters were used, respectively.

To investigate guided-mode resonances supported by the grating structure, a free-space reflectance measurement (Figure 2A) was performed. A broadband supercontinuum laser (SuperK, NKT Photonics) served as the light source, while an optical spectrum analyzer (OSA, ANDOR AQ-6315E) detected the signal. The reflectance was averaged over 10 scans, with a gold mirror as the reference. A 900 nm short-pass filter (FESH0900, Thorlabs) prevented unwanted heating.

### 3.7 Functionalization of the nanostructure

Attachment of the probe to the surface of the array compartment is performed via functionalizing the surface of the nanostructure (Figure S5). The probe was tethered to the array’s nanostructured surface by first introducing reactive aldehyde groups onto the SiNx. Although the native SiNx exposes silanol (Si–OH) and amine/imine (Si–NHC:, SiC:NH) moieties, a brief oxygen-plasma treatment efficiently generates additional surface hydroxyls. The nanostructure-bearing silicon nitride layer was then sequentially cleaned with acetone and RNase⍰free water, dried, and subjected to 5 minutes of oxygen plasma. It was immediately immersed in a 5 % (3⍰Aminopropyl) triethoxysilane solution for 12 hours at ambient temperature, rinsed, dried, and cured at 100 °C for 2 hours. Next, the surface was treated with 5 % glutaraldehyde for 2 hours at room temperature, washed and dried, yielding an aldehyde-activated interface. Probes bearing a C6⍰amino linker were then spotted (1 µL of 100 µM) onto their designated nanostructure sites. To suppress non-specific adsorption, the amplification chamber (C6) was finally treated with 2 % BSA in PBS, followed by a rinse with RNase⍰free water.

## 3. Results and discussion

### 3.1 Nanostructure-enhanced photoluminescence

To evaluate photoluminescence enhancement with the aid of the SiNx grating, we first measured and subtracted the intrinsic photoluminescence (autofluorescence) from both patterned and flat samples as background. We then quantified the effect of the nanostructured substrate by immobilizing 0.1 μM of Cy5 directly on both patterned and flat surfaces (without amplification). The PL spectra, shown in FigureC:2C, exhibited enhanced fluorescence with two distinct peaks centered at 640.3C:nm and 748.4C:nm. We emphasize that we used unpolarized light for excitation and detection, and that resonances from both TE- and TM-polarization can contribute to enhancing the emission.

To determine their origin, we experimentally evaluated the reflection spectra of the fabricated structure in aqueous solution, confirming that the first resonance at 640.3C:nm aligns with the TE-polarized guided-mode resonance (the blue line in Figure 2D), while the second is for the TM guided-mode resonance. The measured reflection spectra are further confirmed by numerical simulations (Figure 2D). Compared to reflectance spectra, the PL resonances appear broadened and shifted, which can be attributed to the presence of the dye with an intrinsic absorption feature. As evident from the PL spectra, the PL intensity near the resonance of 640.3C:nm provides significantly stronger signal enhancement than 748.4 nm. This observation is consistent with our numerical calculations, where this TE-polarized guided-mode exhibited both higher field intensity and broader spatial extension into the surrounding medium, favorable properties for enhancing light–matter interactions. Therefore, we focus our analysis on this resonance.

We further evaluated the fluorescence enhancement factor as the ratio of the net photoluminescence intensity from the nanostructured surface to that from the flat surface. Intensities were integrated over the 630–650C:nm spectral range, associated with the TE guided-mode resonance. The measured enhancement factor reached 6.8 (FigureC:2C), confirming the advantages of using the designed nanostructure in amplifying fluorescence via resonant enhancement. Enhancement factors (Figure 2F) were also measured at other Cy5 concentrations (0.01, 0.1, and 1C:μM), showing similar values except at 1C:μM. We emphasize that the enhancement factor is mainly determined by the enhanced optical field governed by the nanostructure. The small variation of the enhancement factor (Figure 2F) contributed to the inhomogeneous distribution of Cy5, and a slight reduction was likely a result of surface saturation or complete Cy5 coverage on both flat and nanostructured substrates, which impedes the nanostructure’s full enhancement potential. However, notably, this finding highlights the pronounced benefit of nanostructured platforms for detecting low-abundance fluorophores (i.e., low target copy numbers).

### 3.2 Bubble-free and valve-less system

Since our goal of the suggested system is to simplify the microfluidic system without compromising any step of purification and amplification of PCR assay, we propose a vertically operated chip, wherein the fluid naturally remains at the bottom of the chamber due to gravity (Figure 1B and Video S2A). Additionally, Video S2A shows that bubbles are not created during the filling process of PCR chamber (C6) even with intermittent filling that involve air segments. The tendency of the fluid to remain in the bottom eliminates a typical and troublesome issue with bubble formation during the PCR cycling. Unlike numerous works that suggest complicated designs, including multilayer-layer^11^ and bubble trap,^8^ the vertical chip eliminates any generating bubbles instantly to the top of the PCR (Video S2B). Such instant bubble transfer allows a free-bubble system that enables homogeneous reaction and continuous contact of the solid array spots, resulting in a reliable and efficient PCR. In all the previous suggested solid-phase PCR, a mechanism to keep constant contact between the solid surface in contact with liquid phase is absent, rising questions about the reliability of solid-phase PCR.

For fluid control, we suggested a valveless system to maintain a simple orchestration of the chip component. A simple valving system is a critical component to reduce the cost of the total system, wherein the valve manipulates the addition of several reagents for the assay. For simplicity, we designed an on-chip valve with minimum parts. The suggested valve system operates through the two parts of the chip, the inner rotational disc and the outer fixed frame (inset of Figure 1A). The inner disc and the outer frame contain microchannels to connect the reagent chambers. In normally closed status of the valve, the microchannels are misaligned, hence no flow is passed through the microchannels. Once the inner disc is rotated to a specific degree by an aid of an external stepper motor, two microchannels align with each other to allow fluid motion between two points (Video S1A). The vertical chip concept also provides a simple metering system where access fluid is removed when it reaches the scape channel level. The inner disc is made from rigid plastic with a diameter (28.35 mm) slightly bigger than the flexible host frame (27 mm). Hence, the outer flexible frame provides material-tightness on the inner disc due to the diameter mismatch. Such tightness prevents leakage between the interface of the inner disc and the outer frame. The optimal diameter of the inner rigid disc is to be 5% bigger than the diameter of the outer diameter. Smaller diameter dimensions (1-3%) result in a leakage among the chip part while bigger diameter dimensions (>7%) result in a difficulty rotating the inner disc due to the excessive tightness from the outer flexible frame. Although power free multi-valve^24^ and rotary valve chip - Fluidic 155 (from the Microfluidic ChipShop Inc.) suggested rotational valve systems, their system requires multilayers with a layer addition of silicone oil to fill the gap between the layers to prevent inner device leakage increasing the complexity and the cost of fabrication and reducing the robustness of the chip. Our suggested valve system is simpler, and it prevents the leakage by its own structure, while the valve system is a part of the chip itself.

### 3.3 System workflow

To facilitate nucleic acid amplification via PCR, a preliminary nucleic acid purification step is essential. In this study, we developed an integrated microfluidic chip in which all requisite purification and amplification reagents are preloaded into discrete chambers (denoted C1– C7) (Figure 3). Specifically, chamber 1 (C1) contains the lysis buffer/sample, C2 and C3 contain wash buffers 1 and 2, respectively, C4 holds the elution buffer, and C5 contains the RT-PCR reaction mixture. The base of the chip comprises both a PCR reaction chamber and a waste reservoir. A silica matrix, embedded within a precisely fabricated cavity on the inner rotating disc, serves as the solid-phase extraction medium for RNA capture (Figure S3C). During assay operation, reagents are vertically loaded into their respective chambers by the user (Video S1A) as 70 µL of lysis buffer, 70 µL of Wash 1, 70 µL of Wash 2 and 20 µL of elution buffer. The inner disc then undergoes a preprogrammed rotational sequence, aligning microfluidic channels between the disc and the outer chip frame. Simultaneously, an air vacuum is applied at the distal end of the channels to drive fluid flow. The purification sequence (Figure 3A) proceeds as follows: In Step 1, the lysed sample from C1 is introduced at a controlled flow rate (~25 μL/min) into the silica matrix, where RNA is selectively retained. In Step 2, the disc is rotated 8° counterclockwise (CCW) to align C2 with the matrix, delivering wash buffer 1 at a similar flow rate to remove proteins and cellular debris. Step 3 involves a subsequent 8° CCW rotation to introduce wash buffer 2 from C3 for further purification. Waste fluids from steps 1–3 are directed into the designated waste chamber (C7). In Step 4, a 10° CCW rotation aligns the microchannel to bring the elution buffer in C4 with the silica matrix, releasing the captured RNA into the amplification chamber. Subsequently, the disc is rotated once more (30° CW) to bring the RT-PCR mixture from C5 to the eluted RNA in C6, initiating the amplification process. Finally, the motor rotates 10° CW to seal the C6 for RT-PCR process.

**Figure 3.**
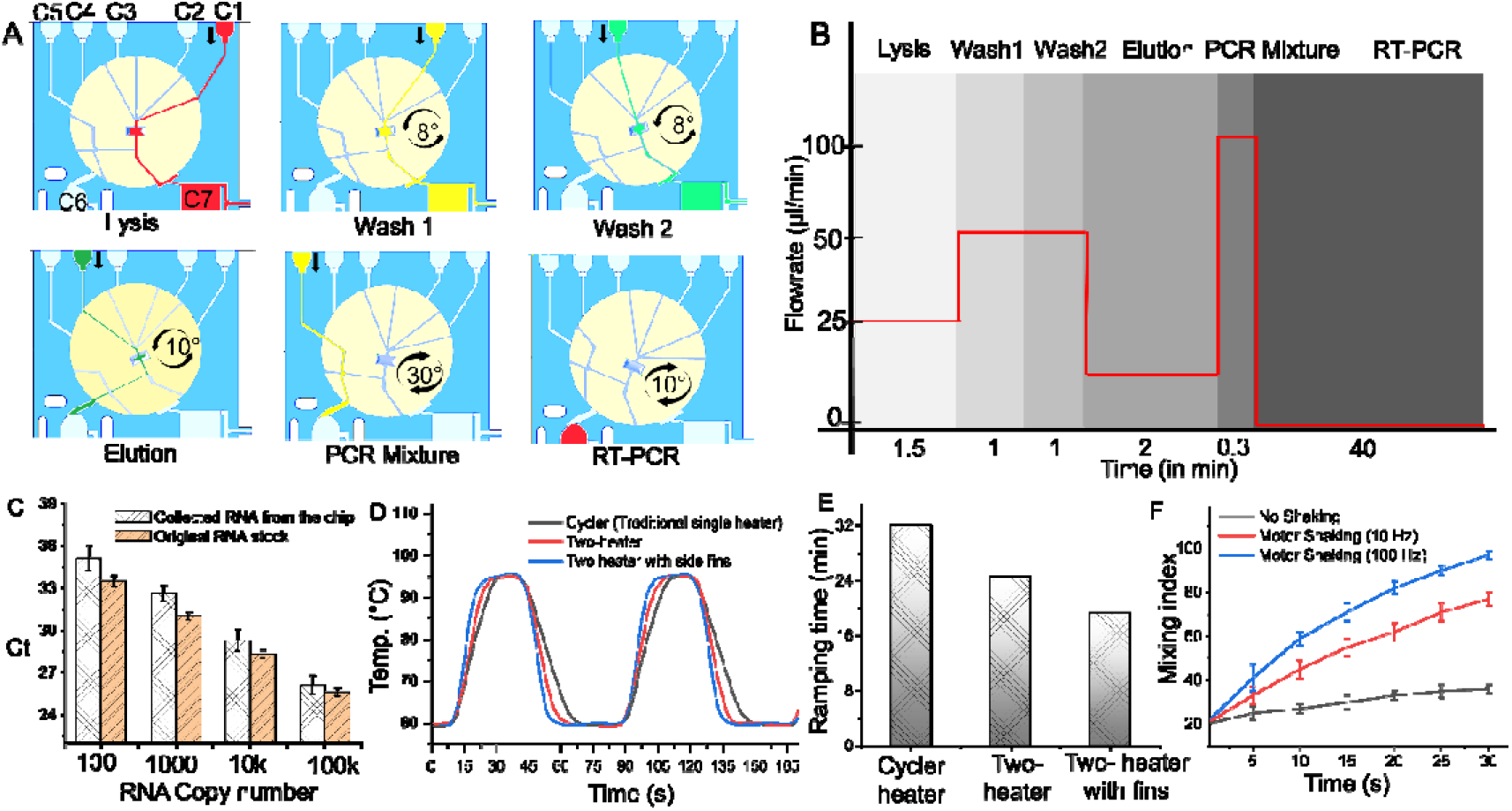
: The workflow of the system. A) step by step of the PCR assay. The lysed sample is enabled by creating a vacuum inside the microchannels forcing the liquid to pass through the silica matrix. Then the inner disc is rotated 8, 8, 10, 30 and 10 degrees sequentially to align microchannels for the addition of the wash 1, wash 2, elution, RT-PCR mixture and sealing, respectively. B) The total time of each assay step corresponds to the flow rate. 6 min for nucleic acid (RNA) purification and 40 min for the solid phase RT-PCR. C) the evaluation of RNA purification on the chip. The chip purifies the RNA with 55% efficiency. D) Thermal performance of two heaters. The two heater metal fins on the side of the PCR chamber provided a faster heating and cooling system since waiting time for the heater to reach the intended temperature is eliminated. E) The total ramping time of the heating system. F) Mixing index with motor shaking.

### 3.4 PCR Process

Once the RT-PCR Mixture and RNA combined in chamber C6, mixing is facilitated via mechanical agitation (Video S2) driven by a fast forwarded and backward motion of an external stepper motor (Stepper Motor 2) mounted on the rear side of the chip. Reverse transcription (RT) is conducted at 50C:°C for 5 minutes to convert RNA into complementary DNA (cDNA), followed by 40 thermal cycles for PCR amplification. To enable rapid and energy-efficient thermocycling, two independent heaters, heater 1 (60C:°C) and heater 2 (95C:°C), were employed (Figure S1B). Rather than ramping temperatures using a single heater, the heaters are alternated beneath the PCR chamber at 0.2-second intervals via the same stepper motor. This dual-heater configuration substantially reduces thermal ramping time by 13 min (Figure 3B, 3D and 3E), eliminates the need for active cooling systems (e.g., fans or heat sinks), and reduces the overall cost by obviating complex thermal controllers, since both heaters operate at constant temperatures. Moreover, the chip’s vertical orientation ensures that the PCR solution consistently remains at the bottom of chamber C6 during cycling. This gravity-assisted configuration eliminates the need for complex chamber sealing mechanisms and minimizes bubble formation, which are common issues in multilayered or sophisticated microfluidic platforms. Although the overall RT-PCR assay time (~45min) remains longer than that of some emerging microfluidic platforms, which is primarily attributed to the use of relatively large reaction volumes (40 μL), the simplicity, scalability, and reduced engineering complexity of the proposed system, particularly the elimination of sealing and multilayer alignment, renders the system a highly viable and practical solution for integrated nucleic acid multiplexed testing. Importantly, the system didn’t compromise any purification steps nor denature (8 s) or extension time (22 s) of the PCR process.

### 3.5 Solid phase PCR on nanostructure

In SP-PCR, the free DNA amplicons attached specifically to the immobilized probes on the solid support. Through a hybridization of newly synthesized amplicons, via LP-PCR, to nested surface-tethered probes, followed by DNA-polymerase-mediated extension of fully matched probes (Figure 4A). To validate the system’s performance, an initial assay targeting SARS-CoV-2 was performed using LP-PCR (without the nanostructured array in C6) and SYBR Green fluorescence detection, which achieved a sensitivity level of 5 copies/reaction (Figure 4B) and a PCR efficiency of 94.8% (Figure S6). Next, for SP-PCR system with nanostructure, we tested the effect of NS compared to the flat substrate with a single viral target (SARS-CoV-2 probe) (Figure 4C-D). The silicon nitride substrate was inserted beneath the PCR chamber (C6), as sim card (Figure S2), with two zones; a zone with an array (3×1) of nanostructures and a zone with flat substrate (Figure 4C). The SARS-CoV-2 probe was immobilized on both zones. At the surface of the NS, the product of the RT-PCR is attached specifically to the target-specific probe. In chamber C6, the setup was loaded with Cy5-labeled forward primers, reverse primers, and DNA produced via reverse transcription of RNA, allowing two amplification processes to occur simultaneously: conventional liquid-phase PCR (LP-PCR) in the solution and solid-phase PCR (SP-PCR) on the nanostructured substrate surface. Following the initial DNA denaturation at 95°C, the resulting single-stranded DNA (ssDNA) amplicons either hybridized with their complementary probes on the solid-phase array, triggering fluorescent labeling and probe extension, or proceeded with standard annealing in LP-PCR at 60°C. At the end of RT-PCR, the PL intensity of the sim was evaluated through integration in a wavelength region 630-650 nm. Figure 4D displays the fluorescence intensity of the solid-phase array at the end of the RT-PCR process (cycle # 37), with the intensity directly reflecting the concentration of amplicons in the solid phases. For the single target (SARS-CoV-2), the integrated system achieved a LOD of 10 copies/reaction and 100 copies/reaction for nanostructures and flat zones, respectively. Hence, integration of the nanostructure with the total system of RNA purification and amplification of the vertical chip guarantees a reliable contact with PCR reagent with the solid surface and assures a clear and distinguishable fluorescent signal intensity from background or possible false positive cases. Nanostructured surfaces significantly enhance fluorescence signals primarily by amplifying light–matter interactions through resonantly enhanced electric fields that extend into the surrounding medium, enabling efficient coupling with nearby analyte molecules. The unique features of the nanostructured surfaces demonstrate the prospects for highly sensitive detection in biosensor applications.^14^

**Figure 4.**
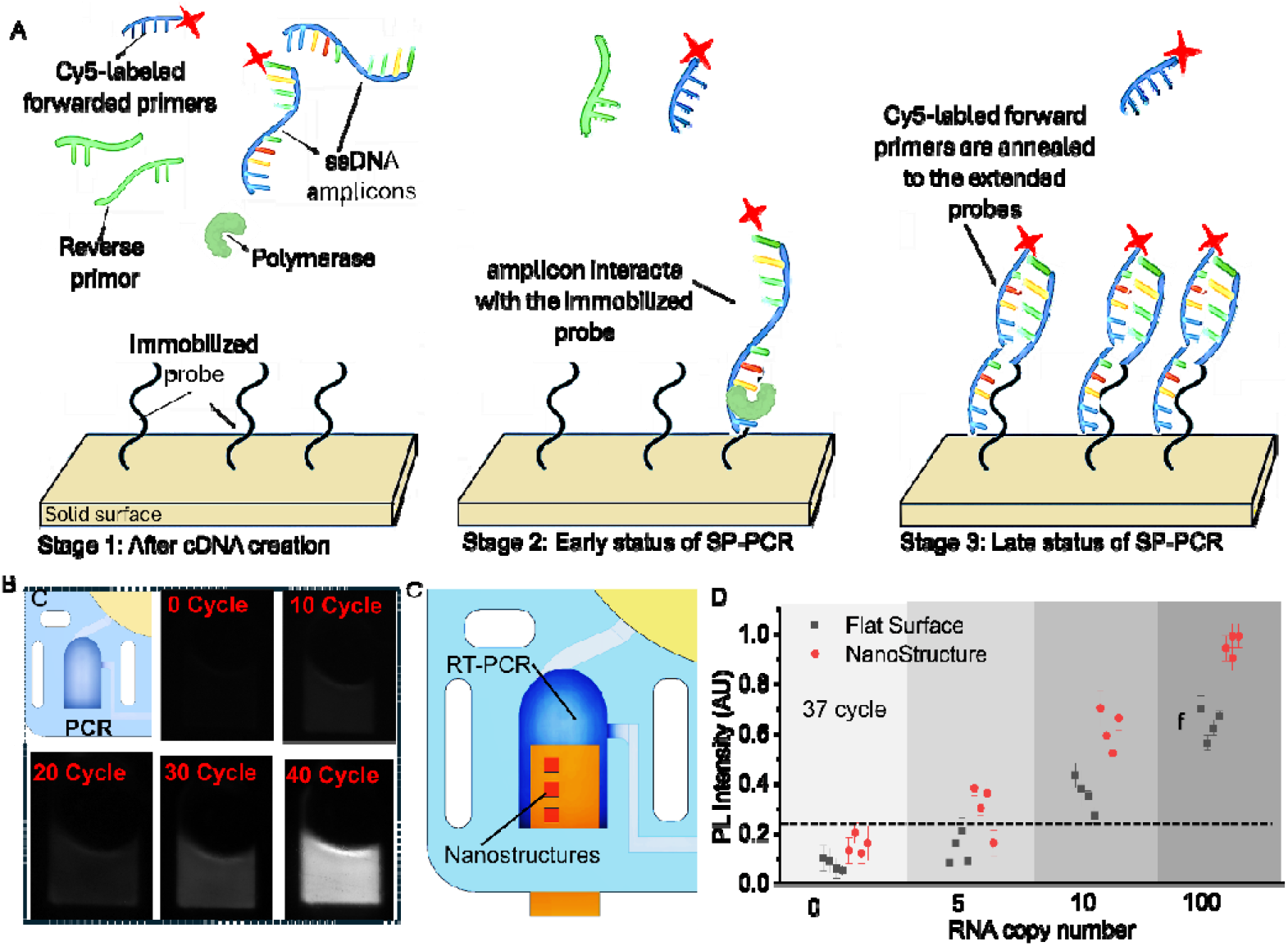
Characterization of SP-PCR assay. (B) The concept of solid phase PCR. In stage 1, after denaturing the cDNA into single stranded DNA (ssDNA) at 95°C, the ssDNA amplicons and PCR components are ready in the liquid phase above the immobilized probe on the solid surface. In stage 2, the newly amplified PCR amplicons in the liquid phase interact with the nested probes immobilized on the solid surface, wherein the matched probes are extended by the polymerase. In stage 3, as the PCR cycles continue, the Cy5-labled forward primers are annealed to the extended probes while more DNA amplicons are created and serve as new templates for the SP-amplification. (B) Time-lapsed images of the fluorescence (CyberGreen) for the traditional liquid RT-PCR. (C and D) Performance of the SP-PCR at flat surface compared to nanostructure (NS) surface. (D) The PL intensity shows the NS surface outperforms the flat surfaces in sensitivity, wherein a 10 copies/reaction of the viral load was detected. Three spots (islands) of NS were measured for each point (n=3). three

### 3.6 Multiplexed Assay and Assay specificity

By arranging an array of different viral target probes, the platform enables multiplexed detection. To thoroughly assess both the analytical performance and specificity of our multiplexed real⍰time assay, we inserted a NS array (3×2) bearing discrete, high⍰affinity oligonucleotide probes against three respiratory viral pathogens (SARS⍰CoV⍰2, Influenza A (Inf. A.) and Influenza B (Inf. B.) (Figure 5A). Following RNA purification, each target was reverse⍰transcribed and amplified via RT⍰PCR directly on the array. When the three viral RNAs were spiked in the sample simultaneously, post⍰amplification fluorescence signatures read after 37 cycles clearly delineated each virus (1000 copies) spiked into the lysis sample (Figure 5B). While when only SARS⍰CoV⍰2 and Inf. A. were spiked, the probe sites for SARS⍰CoV⍰2 and Inf. A. exhibit robust fluorescence signals, whereas the no⍰template control (NTC) spot of the Inf. B remains quiescent. When only Inf. A. was present, the probe sites for the other two viruses only exhibited signals at background levels. The results demonstrated that 100% specificity was achieved when applying fluorescence reading at a cutoff of cycle 37. We noted that background signals could gradually increase beyond 37 cycles, potentially obscuring low⍰abundance positives. Therefore, we operationally classified any fluorescence emerging after cycle 37 as false positive.

**Figure 5.**
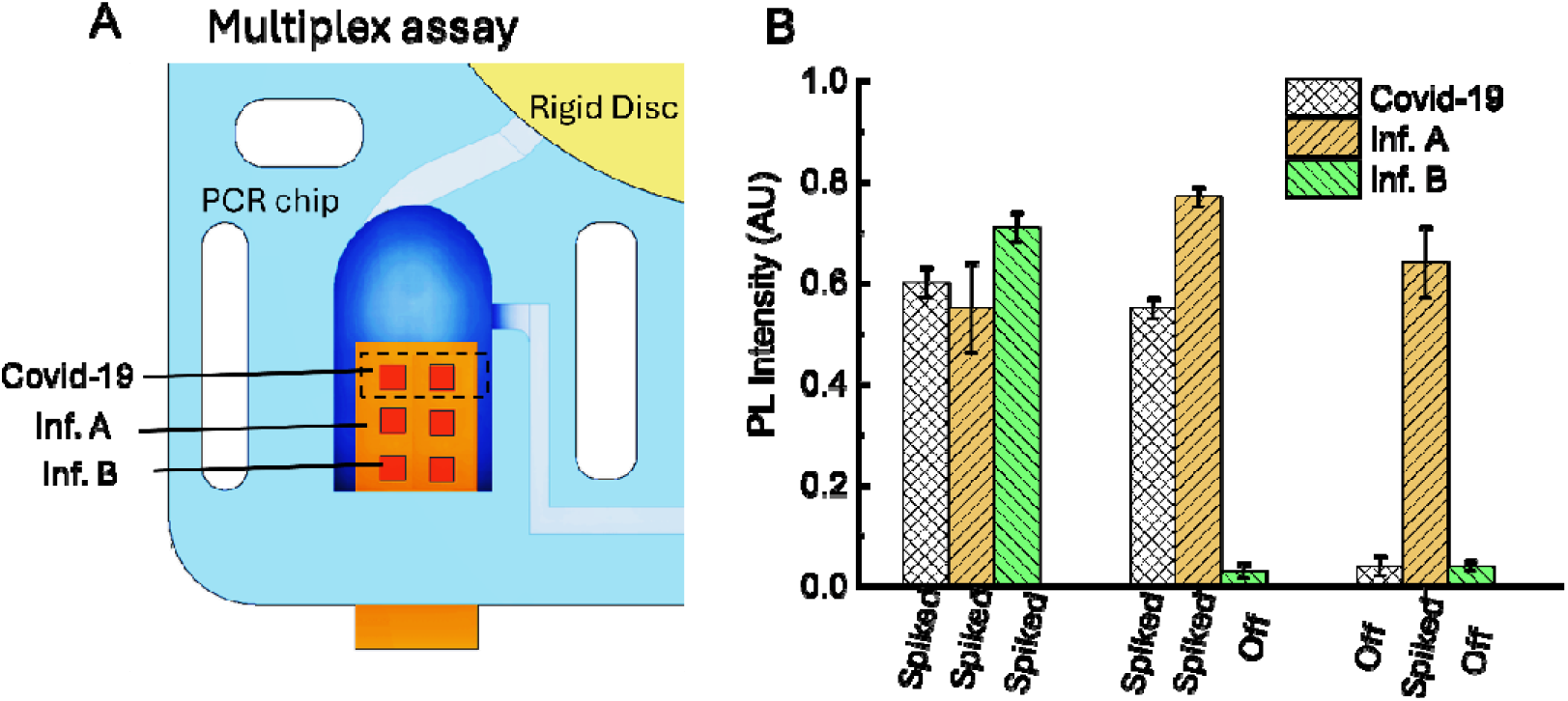
Specificity of the SP-PCR assay. (A) The arrangement of three viruses on the nanostructures inside the PCR chamber. (B) The Specificity test for three viruses: Covid-19, Inf. Inf. A. and Inf. B. spiked sample means viral load (1000 copies) was added to the lysis chamber, while ‘off’ indicates no spiked viral load (0 copy). (n=3).

Previously, it was a concern that the nanostructures may yield elevated background signals in negative samples when compared to planar surfaces of identical material composition,^22^ possibly due to enhanced nonspecific adsorption of fluorescent molecules, a consequence of the high surface energy and complex topography of nanostructured substrates. In our system, we provided adequate surface passivation procedures to prevent retention of fluorescent probes, hence the non-specific binding in the absence of target analytes was minimized.

## 5. Conclusion

In summary, we have demonstrated a fully integrated, valveless microfluidic platform that seamlessly couples rapid RNA extraction, reverse transcription–PCR, and solid-phase amplification on an all-dielectric nanostructured metasurface. Our two-part, vertically oriented chip exploits precise rotational alignment to effect reagent delivery without active valves or pumps, while a dual-heater configuration achieves sub-second thermal cycling without complex cooling systems. The incorporation of a silicon-nitride metasurface beneath the PCR chamber affords photonic-resonance⍰mediated fluorescence enhancement, obviating the need for multiple spectral channels and enabling multiplexed detection with a single fluorophore. This streamlined design achieves a 45⍰minute sample⍰to⍰answer turnaround and a limit of detection of ten RNA copies per reaction for SARS⍰CoV⍰2, underscoring the system’s sensitivity, robustness, and potential for point⍰of⍰care deployment.

Looking ahead, this scalable, low⍰cost architecture holds promise for broadening the scope of molecular diagnostics in both clinical and field settings. By expanding the repertoire of immobilized probes, the platform can be readily adapted for simultaneous surveillance of emerging infectious agents or genetic biomarkers, while its modular fabrication lends itself to high⍰throughput manufacturing. Future refinements, such as integration with portable optical readers, incorporation of automated data analytics, and the development of disposable cartridges, will further enhance usability and accessibility. Collectively, our work establishes a versatile lab⍰on⍰a⍰chip framework that paves the way for personalized medicine, real⍰time pathogen monitoring, and decentralized diagnostic networks.

## Data Availability

All data produced in the present study are available upon reasonable request to the authors

## Acknowledgements

This work was supported by NovoNordisk Foundation, Exploratory Interdisciplinary Synergy Programme, Grant no. NNF21OC0070706. The authors gratefully acknowledge Assoc. Prof. Haiyan Ou and Dr. Osamu Takayama for providing access to their experimental facilities, which were essential for this work

