## Supplementary for "Point-of-Care Solid-Phase PCR in Vertical Microfluidic Chip Integrated with All-Dielectric Nanostructured Metasurface for Highly -Sensitive, Multiplexed Pathogen Detection"

**for**

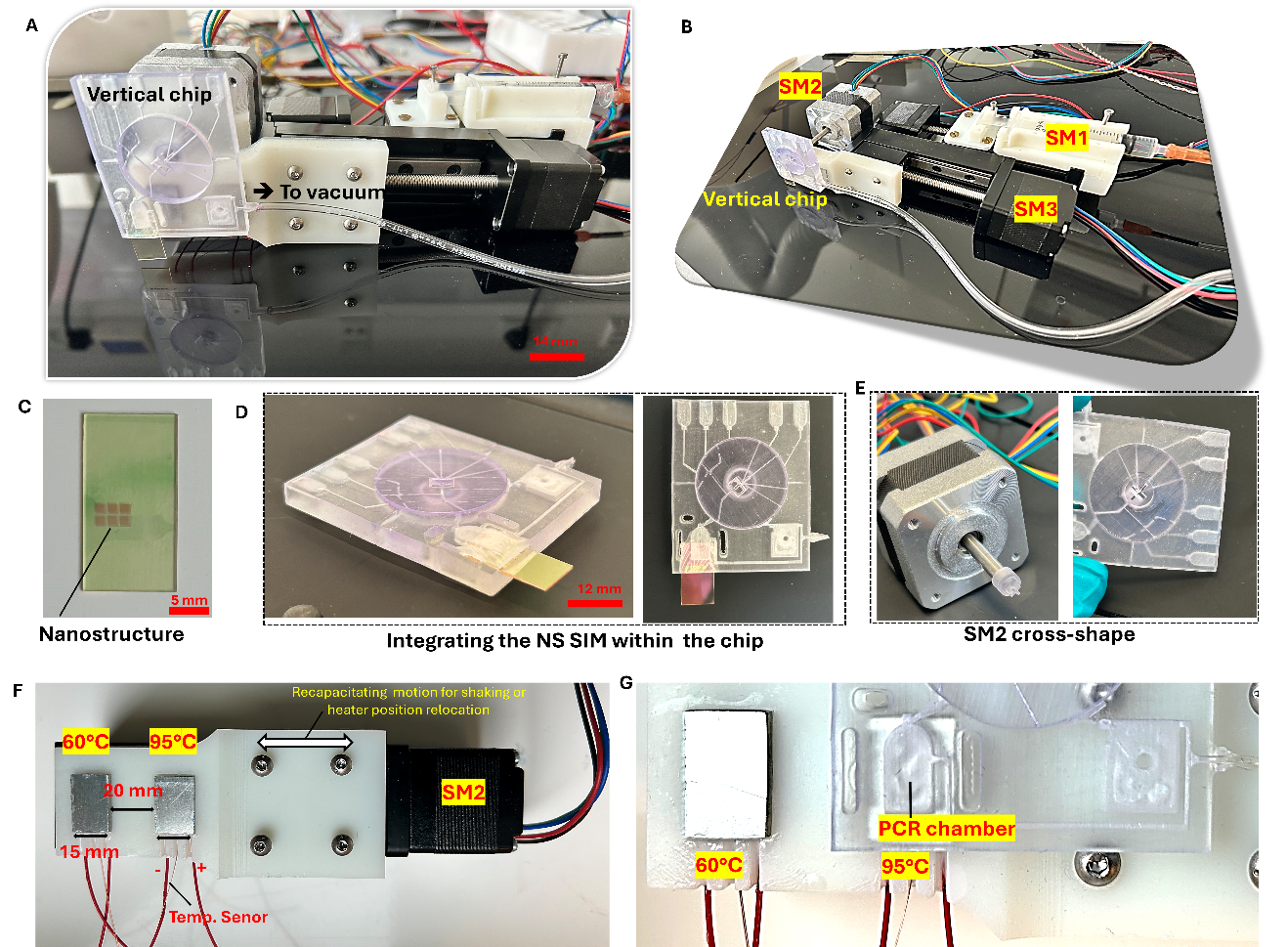

**Figure S1.** Total system. and B. the side and the isometric view of the total system. C. real image of nanostructure islands after fabrication D. Phone-like sim nanostructure car inserted inside the vertical chip. E. the rotational motor. F. the heater system includes two heaters at different temperatures. G. PCR chamber (C6) position**.**

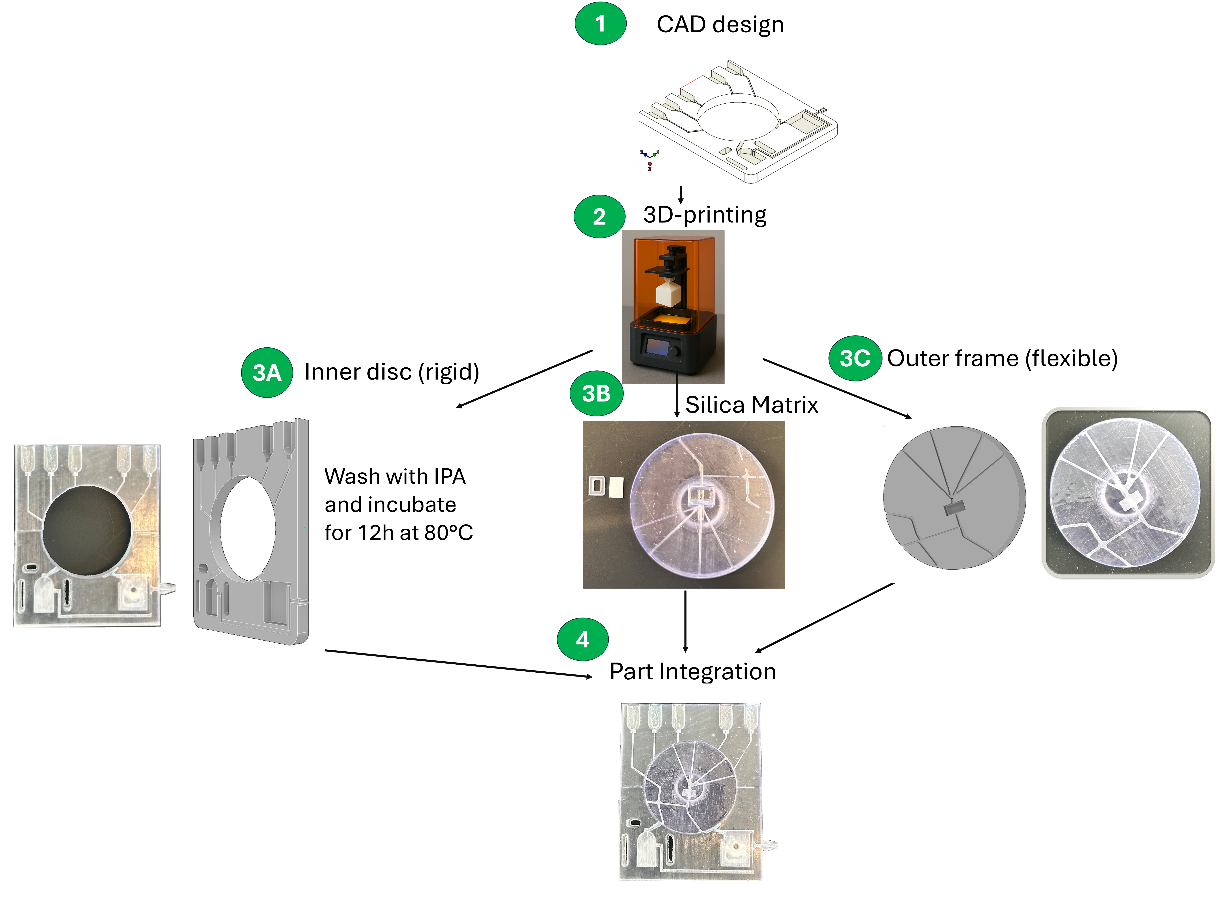

**Figure S2.** System design, fabrication and integration.

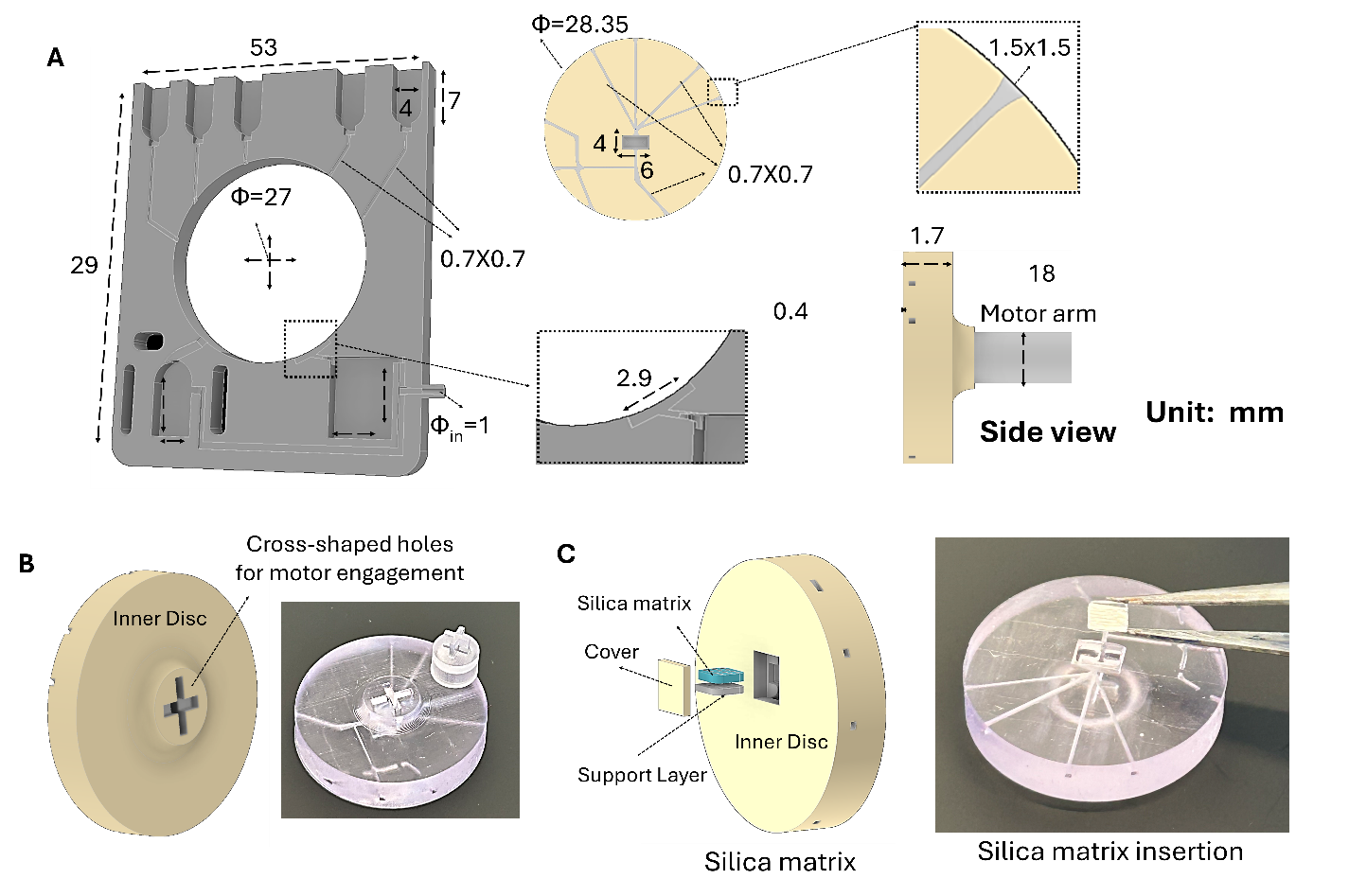

**Figure S3.** System dimension and silicon matrix integration. A. Dimension, in mm, for the outer frame and inner disc. B. The cross-section shape of the back of the inner disc for motor engagement with a real image. C. the silica matrix integration with inner disc.

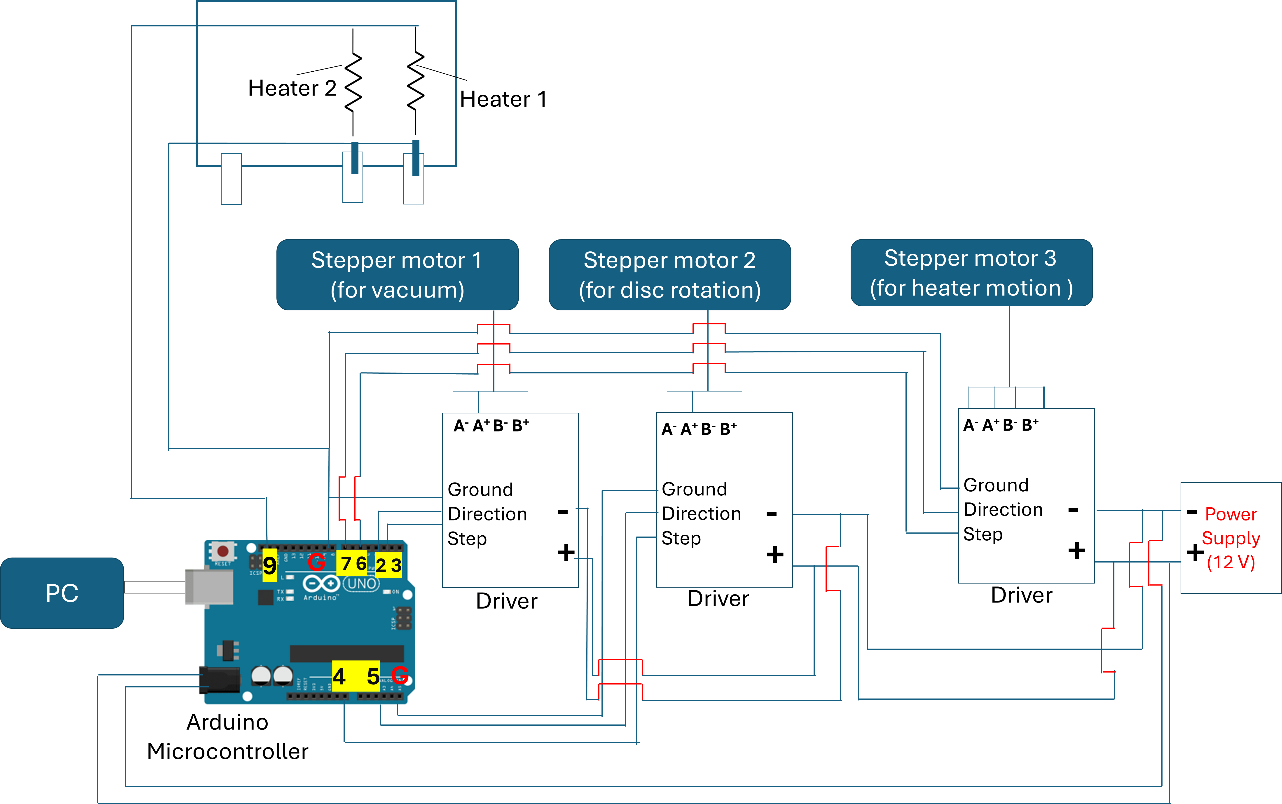

**Figure S4.** Schematic block diagram for Arduino, stepper motors and heaters. The motors are directly connected to motor drivers, which amplifies the low-current to drive the motors. To regulate the motor’s motion, the motor driver is connected to the control circuit (Arduino Microcontroller) through 3 ports: direction, steps and ground. The positive heater is connected to gate 9. Then, code is uploaded into the Microcontroller, which is connected to a PC. The motor driver is powered by aa external power supply and the microcontroller is powered from its USB connection to the PC.

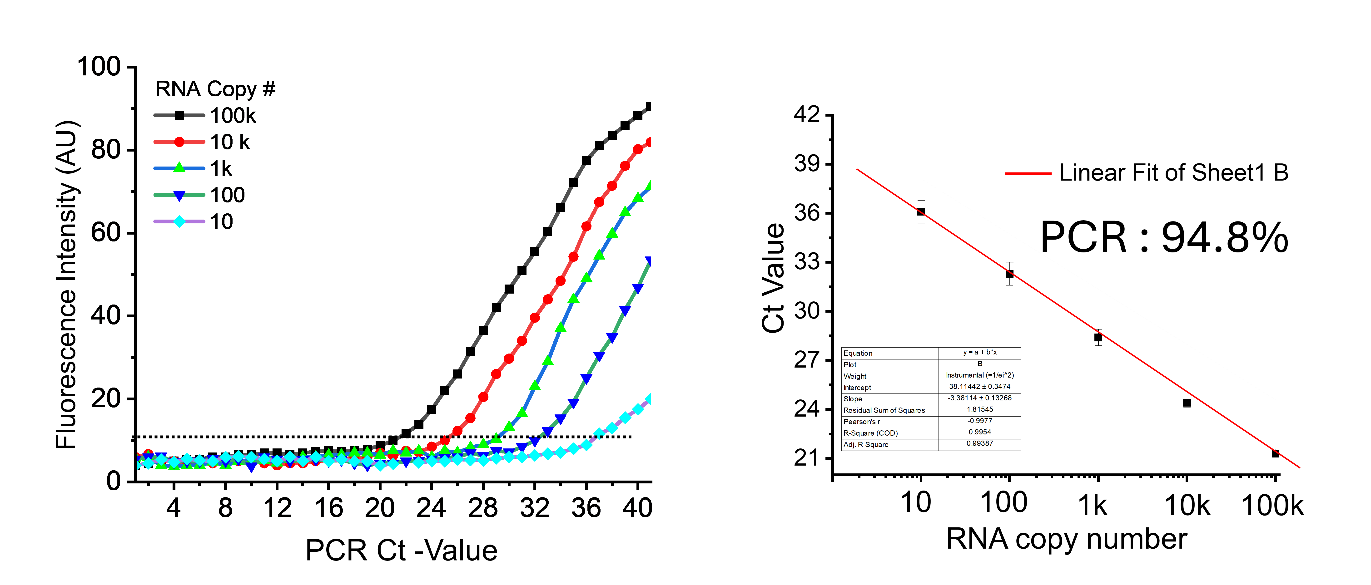

**Figure S6.** A. The Liquid phase RT-PCR performance. A. Real-time singles of the amplified PCR products using SYBR Green fluorescent. B. the PCR efficiency of the liquid phase.

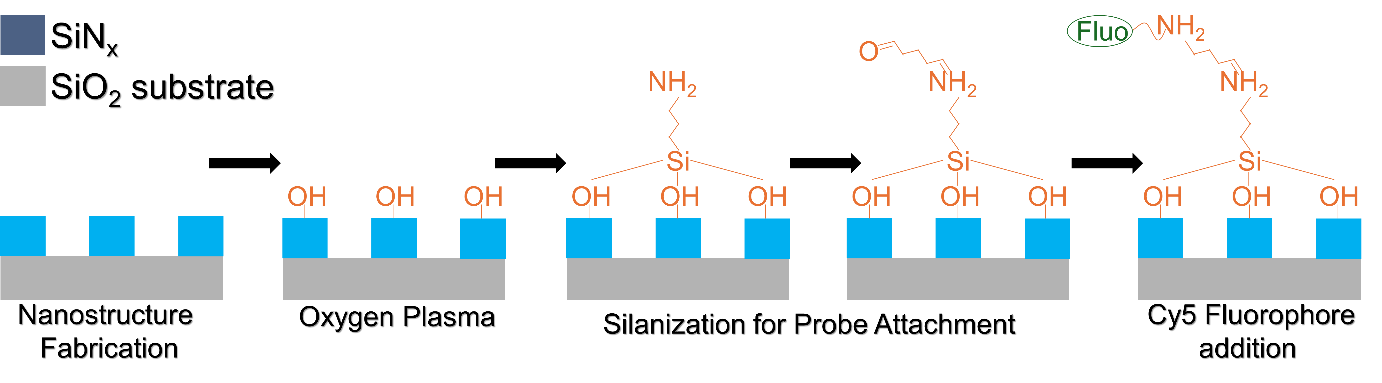

**Figure S6.** Functionalization of the nanostructure

**Estimating Solid-Phase PCR Efficiency (Non-Real-Time Endpoint Measurement)**

The SP-PCR efficiency of both zones resulted in is calculated to be 58.5 % (details in calculation in the supplementary information). The lower amplification efficiency of the SP-PCR (58.5 %) compared to LP-PCR (94.8%) is likely due to steric hindrance, limited molecular diffusion, and surface binding effect. The amplification on solid supports often plateaued earlier, likely due to local reagent depletion. Below are the details for calculating the Solid-Phase PCR efficiency.

Since we did not perform real-time monitoring (no Ct values or exponential amplification curves), traditional qPCR efficiency calculations were not applicable for the solid phase tests. Instead, we estimated solid-phase PCR efficiency based on end-point measurements using the following approach:

PCR efficiency (E) represented the fold increase in DNA quantity per cycle. In ideal solution, phase PCR, 100% efficiency corresponded to a doubling of DNA after each cycle. In our solid-phase system without real-time data, we measured the DNA quantity at the end of the amplification process and back-calculated the efficiency using the known initial quantity of template and the number of cycles performed. In the endpoint PCR, where only the final amplified product is measured, the approach differs. Now, to estimate PCR efficiency from endpoint PCR, we prepared a serial dilution series of the RNA template, typically in 10-fold steps (100K, 10k, 1k, 100 and 10 copies/reaction). Then, we performed RT-PCR amplification for each dilution under identical cycling conditions. Next, we quantified the final PCR products by electrophoresis via resolving the PCR products on an agarose gel and then stained the gels using ethidium bromide or SYBR Safe. We imaged the gels and measured the band intensities using ImageJ or GelAnalyzer software. Finally, we constructed a standard curve by plotting X-axis: logarithm of the initial DNA template concentration and Y-axis: logarithm of the measured product quantity (based on band intensity) and we determined the slope of the resulting linear regression line.

To calculate the amplification efficiency (E) for liquid and solid phase, we used the formula:

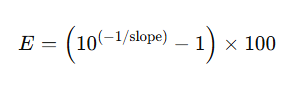

Ideally, a PCR with 100% efficiency would exhibit a doubling of the product in each cycle. However, in endpoint PCR, efficiency estimation is less precise due to the plateau phase, during which reagent depletion and product accumulation slows the amplification rate. Therefore, for improved accuracy, we limited the number of PCR cycles to maintain amplification within the exponential phase.

**Logistic curve parameters**

To fit experimental data the Logistic function was used [1]:

*y=A2+(A1-A2)/(1+(x/x0)^p),*

here x - is the number of copies, and A1, A2, x0 and p are fitting parameters. The obtained fitted parameters are presented in Table S1.

**Table S1.** Fitted parameters obtained using the Logistic function.

| **Structures** | **Target analyte** | **A1** | **A2** | **x0** | **p** | **R2** |
| --- | --- | --- | --- | --- | --- | --- |
| Nanostructures | Covid | 13490 | 1.62E8 | 3.05E13 | 0.22 | 0.932 |
|  | Influenza A | 2550 | 9.53E7 | 2.16E13 | 0.19 | 0.943 |
|  | Influenza B | 12549 | 2.60E6 | 3987 | 0.30 | 0.987 |
| Flat surface | Covid | 19029 | 1.39E6 | 6.07E5 | 0.43 | 0.998 |
|  | Influenza A | 20853 | 4.72E8 | 2.29E9 | 0.59 | 0.991 |
|  | Influenza B | 6337 | 1.45E7 | 1.54E8 | 0.44 | 0.998 |

**Table S2: Reagent for RT-PCR**

| **Liquid phase RT-PCR Solution 20µl (according to the manufacturer)** | | |
| --- | --- | --- |
| *Reagent* | *Amount (µl)* | *Final concentration* |
| 2x SensiFAST mix | 10 |  |
| Reverse Transcriptase | 0.2 |  |
| RNase inhibitor | 0.4 |  |
| Normal Forward 10µM | 0.8 | 0.4µM |
| Normal Reverse 10µM | 0.8 | 0.4µM |
| EvaGreen | 0.5 |  |
| RNA | 1 |  |
| Water | 6.3 |  |

**Table S3: Sequence of the probe and the primers**

| **Primers and probes used for RT-PCR** | | |
| --- | --- | --- |
| **Target** | **Sequence** | *Used concentration* |
| **Forward primers (Cy5-labeled)** | | |
| Covid-19 | Cy5-GATCTCAATGGTAACTGGTATGATTTCGGTG | 10µM |
| Inf. A | Cy5-TAACGGGAAACTATGCAAACTAAGAGG | 10µM |
| Inf. B | Cy5-GGGATAGAGATGGTACACGATGGTG | 10µM |
| Reverse primers | | |
| Covid-19 R | GCCCTGGTCAAGGTTAATATAGGCATTAAC | 10µM |
| Inf. A | GTGTTTCCACAATGTAGGACCATG | 10µM |
| Inf. B | TGTGACAGTGTCCCATAGCAA | 10µM |
| **Probes used for immobilization (C6-attached)** | |  |
| Cy5-probe | C6-GTAATTGATTAGCTTGTCGTTGTGA-Cy5 | 100µM |
| Probe- Covid-19 | C6-GAATCTACAACAGGAACTCCACTACCT | 100µM |
| Probe- Inf. A | C6-CTGTGGAGAGTGATTCACACTCTGGA | 100µM |
| Probe- Inf. B | C6-GGCTGTTGCAGCTGAATGCCAAGT | 100µM |
